# Evaluating Cognitive Impact of Traumatic Brain Injury and Risk for Post-Traumatic Epilepsy

**DOI:** 10.64898/2026.08.30.26361760

**Authors:** Taylor Zink, Henry Noren, Daniel Valdivia, Christine Yohn, Jasdeep Hundal, Spencer Chen, David Scarisbrick, Hai Sun

## Abstract

**Objective:** Post-traumatic epilepsy (PTE) is a common sequela of traumatic brain injury (TBI). Research indicates that individuals with PTE tend to experience greater cognitive difficulties compared to those with TBI alone. However, it is plausible that a distinct cognitive profile exists that distinguishes between TBI cases with and without PTE. We aimed to identify longitudinal changes in cognitive measures among TBI patients to better assess the changes associated with developing PTE.

**Setting:** Outpatient.

**Participants:** Prospective subjects who had suffered TBI within 6 months post-injury (TBI-6M, n=32), retrospective subjects with pre-existing PTE diagnoses (PTE, n=20), and healthy control subjects (HC, n=41).

**Design:** We examined cognitive performance for TBI patients within 6 months post-injury, then again within 12 months (TBI-12M, n=26), and within 18-months (TBI-18M, n=25), and compared this with cognitive performance among HC and PTE.

**Main Measures:** Cognitive tests administered yielded 15 test components for analysis. We utilized linear mixed effects modeling to examine cohort-level differences cognitive function.

**Results:** 11/15 tests showed a significant performance deficit in the PTE subjects compared to HC. TBI-6M was not significantly different from the PTE subjects; with time, 9/15 tests showed some degree of recovery in TBI subjects. Tests for information processing speed/working memory and executive function showed strong recovery (TBI-6M vs. TBI-18M, SDMT written: p<0.0001, SDMT oral and COWAT: p<0.001). Tests for visual attention/working memory also showed a smaller but significant recovery (TBI-18M vs. PTE, p<0.05). By contrast, tests for verbal memory [HVLT-R Delayed Recall] showed chronic impairment in TBI (TBI-18M vs HC, p<0.0001). TBI subjects generally trend towards recovery in cognitive performance post-TBI.

**Conclusions:** Information processing speed/working memory are strong indicators for TBI recovery, while auditory learning/memory shows chronic impairment. The stagnation of recovery in cognitive domains typically characterized by robust recovery may correlate with an elevated risk of developing PTE.

## Introduction

Traumatic brain injury (TBI) is a major cause of death and disability worldwide and is associated with substantial long-term neurological, cognitive, and psychosocial morbidity ^1^. Among the most devastating long-term sequelae of TBI is post-traumatic epilepsy (PTE), an acquired seizure disorder characterized by recurrent, unprovoked seizures occurring more than seven days after the initial injury ^2,3^. The reported incidence of PTE varies widely on injury severity and mechanism, with rates ranging from approximately 3-5% after closed-head injury to over 50% or more after penetrating head injury ^4^. Long-term military cohorts with penetrating injuries have reported PTE prevalence as high as 45-53% ^5^. Greater injury severity, early post-traumatic seizures, intracranial hemorrhage, and temporal lobe injury are recognized risk factors for PTE development ^6,7^.

PTE is particularly devastating because it may impair neurological recovery, worsen functional outcomes, reduce quality of life, and increase long-term cognitive, emotional, and psychosocial burden ^8,9^. Cognitive dysfunction is among the most reported chronic consequences of both TBI and epilepsy, affecting cognitive domains such as attention, memory, executive functioning, processing speed, language, and visuospatial ability ^9,10^. Because both TBI and epilepsy independently contribute to cognitive impairment, patients who develop PTE may experience an even greater cumulative neurocognitive burden ^10^. Although antiseizure medications are commonly prescribed with the aim of reducing early post-traumatic seizures, no established therapy currently prevents epileptogenesis or reliably eliminates downstream PTE development ^11,12^. Because PTE may emerge months to years after the initial injury, identifying early clinical markers of increased risk is a major unmet need ^13^.

While prior work has focused on injury severity, seizure burden, neuroimaging findings, and EEG biomarkers, fewer studies have examined whether longitudinal neurocognitive recovery patterns may help distinguish patients at increased risk for PTE ^14,15^. Reviews have identified fewer than ten studies which examine the relationship between cognitive functioning and PTE, with even fewer of these studies using formal neuropsychological assessments ^10,16^. These studies suggest that patients with PTE experience greater cognitive impairment than patients with TBI alone; however, the specific cognitive domains most strongly associated with epileptogenesis remain poorly understood ^15^. In a large cohort 20-year study of U.S. veterans, cognitive dysfunction was one of the strongest mediating factors of epilepsy risk after TBI identified, suggesting cognitive deficits may not present merely as an outcome of PTE but as a potential intermediary in epileptogenesis ^17^. Another study demonstrated that the burden of seizures was associated with worse global cognition at 3 months post-TBI, but did not track the trajectories of early cognitive impairment as a predictor of PTE ^18^.

Here, we contribute to the limited but growing literature describing longitudinal neurocognitive outcomes following TBI and further characterize neurocognitive performance in patients with established PTE. In this study, we aimed to evaluate longitudinal changes in cognitive assessment performance following TBI across multiple cognitive domains over three timepoints. We also compared these trajectories with the cognitive performance of a cohort of individuals with PTE to identify cognitive patterns that may distinguish PTE from TBI alone and better characterize the additional cognitive burden associated with post-traumatic epilepsy.

## Methods

### Study Participants

All participants provided informed written consent, and the study was approved by the Rutgers eIRB. Three cohorts of participants between 18 and 65 years of age were recruited: healthy controls (HC), individuals with established PTE as a retrospective cohort (PTE), and individuals with recent TBI as a prospective cohort (TBI). Three healthy controls were excluded after screening positive for a prior history of TBI. The PTE cohort included subjects with a history of TBI preceding seizure onset, a diagnosis of PTE by an epileptologist, and video EEG evidence of focal epilepsy.

The prospective TBI cohort included subjects who sustained a moderate-to-severe TBI or complicated mild TBI within 6 months of enrollment. Complicated mild TBI was defined by the presence of a contusion on presenting CT or MRI. Subjects underwent longitudinal data acquisition within 6 months of injury (TBI-6M), with additional assessments approximately 12 and 18 months after injury (TBI-12M and TBI-18M). TBI subjects were excluded for a pre-existing neurological or psychiatric diagnosis, history of substance abuse, prior seizures or stroke, or craniotomy before the index TBI. Across cohorts, subjects whose primary language was not English were excluded to reduce language-related effects on cognitive performance.

### Data Collection

Demographic and medical history information was collected via a customized REDCap questionnaire and electronic medical record review. Collected data included demographics, education and socioeconomic status, seizure and TBI history, medical history, medications, and substance use. For PTE subjects, when medical records were unavailable to verify dates of initial TBI and seizure onset, seizure onset was estimated based on the patient’s reported age at seizure onset and age at initial TBI. Due to missing injury characteristic data, initial injury severity was compared between TBI and PTE cohorts using self-reported loss of consciousness (LOC) and post-traumatic amnesia (PTA) (Supplemental Table 1).

### Cognitive Measures and Scoring

We administered a variety of cognitive assessment measures to cover a wide range of cognitive domains. Cognitive measures administered included the Neuropsychological Assessment Battery (NAB) Digits Span and Dots tests ^19^, Trails Making Test A and B (TMT) ^20^, the Stroop Color and Word Test Adult Version ^21,22^, Symbol Digits Modality Test (SDMT) ^23^, Controlled Oral Word Association Test FAS version (COWAT) ^24^, Brief Visuospatial Memory Test-Revised (BVMT-R) ^25^, and Hopkins Verbal Learning Test-Revised (HVLT-R) (Table 2) ^26,27^. Tests were scored using official manuals for their respective batteries. Assessed cognitive functions for each test were derived from official scoring manuals and previously validated studies ^22,28,29^. Cognitive measures were administered in a standardized order to minimize confounding effect on performance.

### Statistical Analysis

All statistical analyses were performed in RStudio (R version 4.4.2). Fisher’s exact test was used to assess gender, race, ethnicity, SES, and education level across cohorts, and one-way ANOVA was used to assess differences in age (Table 1). Educational attainment was treated as an ordinal variable using a 4-point scale. Pearson correlation was used to assess the relationship between time from injury to first testing session and cognitive measure scores, with no significant relationships observed (Supplemental Table 1).

**Table 1.** Demographic Data. P-values for gender, race, ethnicity, SES, and education level were calculated using Fisher’s exact test. P-value for age were calculated using One-Way ANOVA.

|  | HC | PTE | TBI | p-value |
| --- | --- | --- | --- | --- |
| **Age (avg. ± SD)** | 36.0 ± 13.3 | 38.4 ± 9.95 | 41.1 ± 13.1 | 0.273 |
| **Race (n)** |  |  |  | 0.076 |
| *White* | 25 | 8 | 15 |  |
| *Asian* | 9 | 1 | 4 |  |
| *Black/African American* | 5 | 8 | 8 |  |
| *Not answered/Prefer not to answer* | 2 | 3 | 5 |  |
| **SES (n)** |  |  |  | 0.096 |
| *<\$15,000* | 1 | 4 | 2 | |
| *\$15,000-74,000* | 12 | 6 | 17 | |
| *>\$100,000* | 21 | 7 | 10 | |
| *Unknown or refuse to answer* | 7 | 3 | 3 |  |
| **Gender (n)** |  |  |  | 0.003* |
| *Male* | 18 | 16 | 25 |  |
| *Female* | 23 | 4 | 7 |  |
| **Ethnicity (n)** |  |  |  | 0.079 |
| *Hispanic/Latino* | 4 | 2 | 5 |  |
| *Not Hispanic/Latino* | 36 | 13 | 24 |  |
| *Not answered* | 1 | 5 | 3 |  |
| **Education level (n)** |  |  |  | 0.006* |
| *Less than high school or unknown* | 0 | 1 | 1 |  |
| *High school or GED* | 6 | 6 | 13 |  |
| *Some college or bachelor's degree* | 16 | 11 | 13 |  |
| *Graduate degree or greater* | 19 | 2 | 5 |  |

To examine cognitive measure performance, cohort, age, and educational level were modeled as fixed factors, with subject ID as the random factor in a linear mixed effects (LME) model. The output was analyzed using emmeans to derive estimated means, standard errors, and confidence intervals. Pairwise comparisons were used to assess significance (p ≤ 0.05) between cohorts.

Additionally, raw scores were analyzed using Spearman’s correlation to assess redundancy among cognitive measures. Correlations greater than 0.7 were treated as significant redundancy. To assess whether cognitive measures could be grouped into domains for simplified analysis, we ran two additional LME models as described previously. The first combined measures based on the cognitive function they were designed to assess (theoretical domains): Working Memory, Visual Learning and Memory, Auditory Learning and Memory, Basic Processing Speed, Complex Processing Speed, and Executive Functioning. The second combined measures based on Spearman’s correlation with hierarchical clustering set to four clusters (computed domains): Processing Speed, Audiovisual Memory, Auditory Attention, and Cognitive Flexibility.

## Results

### Participant Characteristics and Injury Severity

We included 41 control, 20 PTE, and 32 TBI subjects (TBI-6M timepoint) in the study (Table 1). At the time of this analysis, 26/32 TBI subjects had completed their second timepoint data acquisition (TBI-12M) and 25/32 had completed their third timepoint data acquisition (TBI-18M). Fisher’s exact test revealed significant differences in gender (p = 0.002), and educational level (p = 0.004) across cohorts. The control cohort was 43.9% male (18/41), compared with 80.0% male in the PTE cohort (16/20) and 78.1% male in the TBI cohort (25/32), consistent with known sex-based differences in TBI epidemiology ^30^. Education level also differed across cohorts. Graduate-level education or higher was present in 46.3% of controls (19/41), compared with 15.6% of TBI subjects (5/32) and 10.0% of PTE subjects (2/20).

To better compare the initial injury severity of our TBI and PTE cohorts, we compared self-reported LOC, anterograde amnesia, and retrograde amnesia. Of the symptoms assessed, LOC greater than 5 minutes was significant between TBI and PTE cohorts (p < 0.05), which on further investigation revealed that the TBI cohort more frequently reported this symptom than PTE. No other symptoms assessed showed significance between cohorts. Additionally, we compared individuals in the PTE cohort missing LOC/PTA data to those with their data via independent t-test, and saw no significant differences in cognitive assessment performance.

### Longitudinal Neurocognitive Outcomes

To evaluate performance on cognitive measures between our cohorts and longitudinal performance in TBI, we fit a linear mixed effects model on cohort, age, educational level, and subject ID, then evaluated the resulting means via Pairwise comparisons. Out of the fifteen subtests that we derived from our battery, eleven of them showed significance (p < 0.05, pairwise comparisons) in scores between PTE and HC, indicating PTE had impairment of a wide variety of neurocognitive functions. No tests showed significance when comparing PTE to TBI-6M, indicating that the initial impairment in cognitive function due to TBI was comparable to the impairment due to PTE. Seven out of fifteen tests showed significance when comparing TBI-6M to HC, indicating a strong initial impairment of cognitive function due to TBI. The COWAT, Stroop Color-Word, SDMT Oral, and SDMT Written tests showed significance when comparing TBI-6M to TBI-18M, indicating a strong recovery trend in the oral/written processing speed, working memory, executive function/letter fluency, and inhibitory executive control/selective attention domains (Figure 1A). The SDMT Oral, SDMT Written, and the Dots subtest of the NAB showed significance when comparing TBI-18M to PTE, indicating recovery of function in the associated cognitive domains. The Stroop Word, Stroop Color, HVLT Trials 1-3 and HVLT Delayed Recall showed significance when comparing TBI-18M to HC, indicating prolonged impairment of cognitive function in verbal processing speed, auditory learning/encoding and memory/retrieval, with HVLT Delayed Recall showing no change in significance level between testing sessions for TBI subjects, indicating permanent loss of function (Figure 1A).

**Figure 1.**
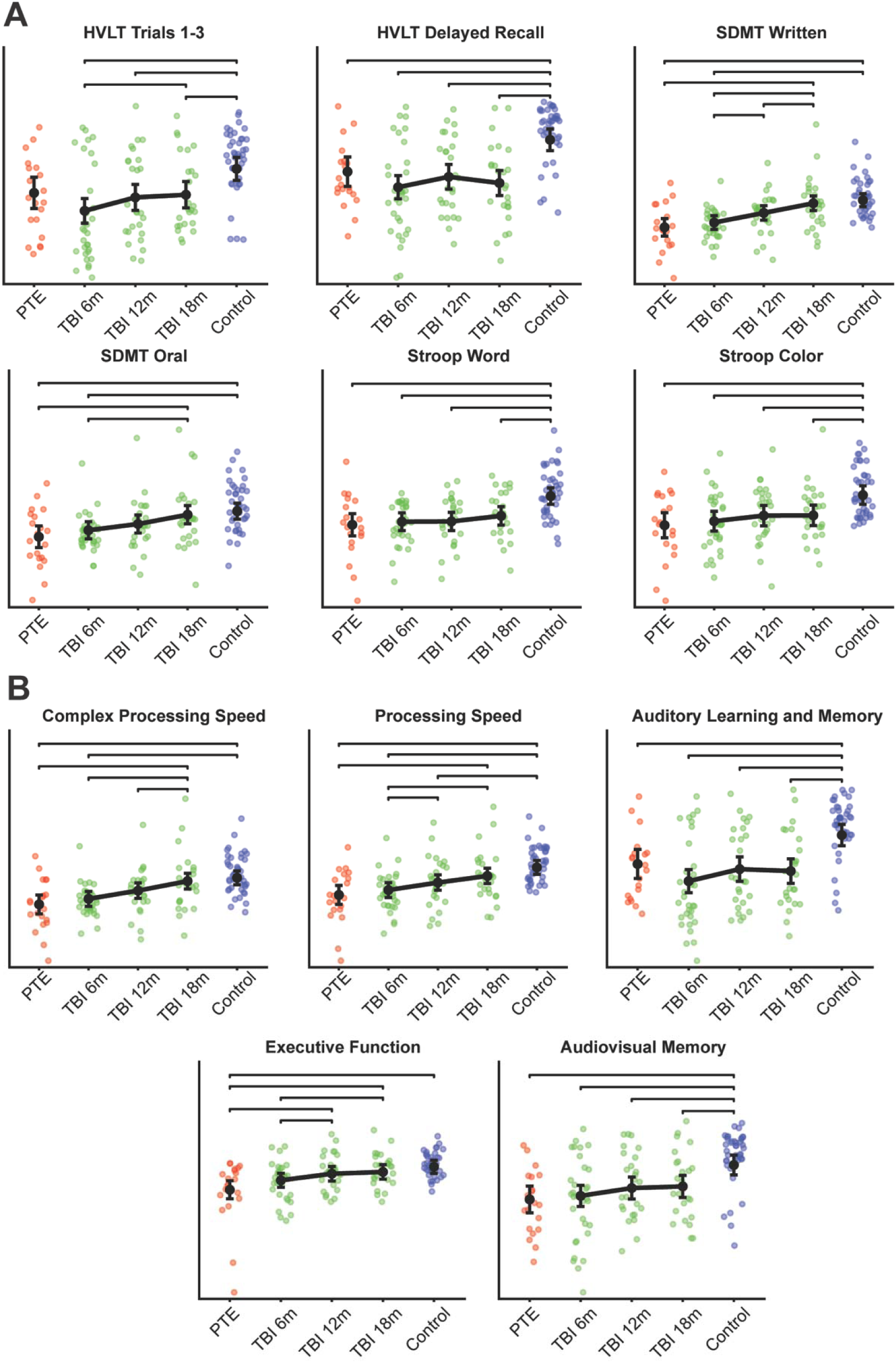
(A) Trends in key cognitive measures. (B) Trends in key domains.

### Cognitive Domain Analyses

The Spearman’s correlation showed a total of five redundancies between cognitive measures (Figure 2). These redundancies were between Stroop Word and Stroop Color, Stroop Color and Stroop Color-Word, SDMT Oral and SDMT Written, BVMT Trials and BVMT Delayed Recall, and HVLT Trials and HVLT Delayed Recall. These redundancies are to be expected, given that the redundancies seen are between measures within the same original battery and are assessing similar cognitive functions (Table 2). Among measures within differing batteries, there is a tendency for a weaker positive correlation that does not reach the level of redundancy.

**Figure 2.**
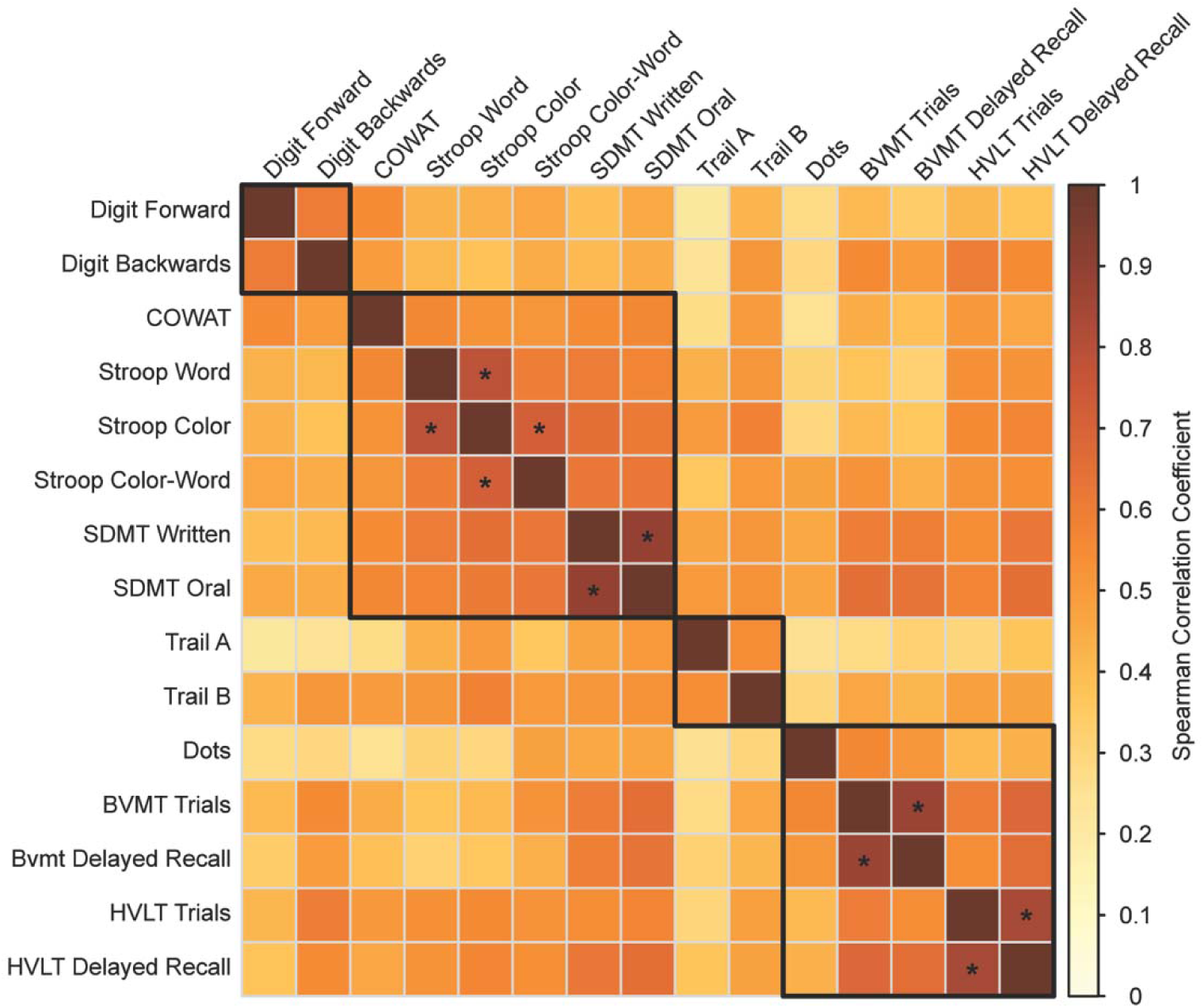
Heatmap that shows the output of Spearman’s correlation of the raw cognitive measure scores across all cohorts. Highlighted black boxes indicate grouping of measures, and values greater than 0.7 (indicated by *) indicate significant redundancy between cognitive measures.

**Table 2.**
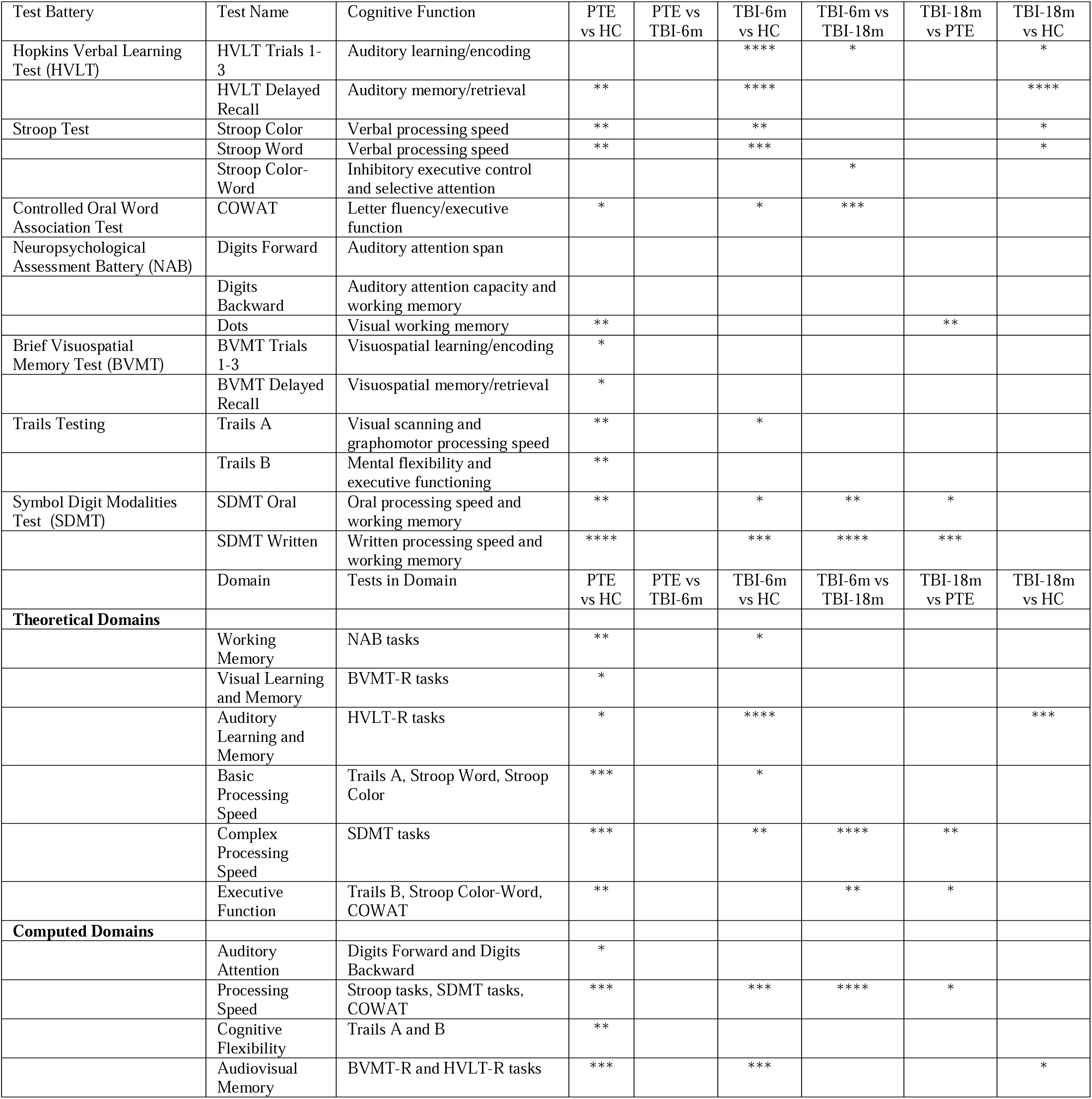
Cognitive Data. P-values of the pairwise comparisons of the means for cohorts derived from the linear mixed effects model. *p < 0.05, **p < 0.01, ***p< 0.001, ****p<0.0001.

In the theoretical domain analyses, all six domains showed significant differences between PTE and HC, indicating widespread impairment of cognitive functioning in PTE (Table 2). Working Memory and Basic Processing Speed showed impairment between HC and TBI-6m, indicating initial impairment of those functional domains post-TBI. Executive Function and Complex Processing Speed showed significant differences between TBI-6m and TBI-18m, as well as between PTE and TBI-18m, indicating significant recovery of these functions in TBI patients (Figure 1B). Auditory Learning and Memory showed significance between HC and TBI-18m, indicating prolonged recovery of function or chronic impairment (Figure 1B).

For the computed domains analysis, as with the theoretical domains, all four domains showed significant differences between PTE and HC, once again indicating the widespread cognitive impairment seen in PTE (Table 2). Processing Speed showed significance between TBI-6m and TBI-18m, as well as significance between PTE and TBI-18m, indicating significant recovery in TBI patients (Figure 1B), while Audiovisual Memory showed significance between HC and TBI-18m, indicating prolonged recovery of function or chronic impairment (Figure 1B).

## Discussion

The current literature has illustrated that PTE leads to impaired recovery of cognitive performance after TBI compared with TBI patients who do not develop PTE ^10^. Our data has shown that the absence of significant differences in cognitive test results between individuals with PTE and those with TBI suggests that the cognitive impairment observed shortly after a TBI is similar to that of PTE. Additionally, the individuals with PTE were, on average, several years post-injury, which, when taking our cognitive measure findings into consideration, suggests that PTE causes an impairment or arrest in cognitive recovery sometime post-TBI (Supplemental Table 1).

### Cognitive Recovery Following TBI

Several prior studies have found that cognitive performance improves in the early recovery stages post-TBI but shows chronic cognitive impairment in the late stages of recovery compared with healthy controls ^31,32^. Our results indicated that oral and written processing speed, along with working memory, exhibited the strongest trends toward recovery among all the measures assessed. Notably, these measures reached a level of non-significance when compared to healthy controls (HC), suggesting a strong potential for functional recovery following TBI. Current literature suggests a long recovery period for generalized cognitive function post-TBI ^33^, but our results suggest that the speed of recovery varies among different cognitive functions. Our results showing a variation of recovery speed are further supported by other studies showing that working memory, executive function, and oral processing speed display a propensity towards recovery after TBI ^34,35^. Different cognitive functions may recover at different rates after TBI because they rely on distinct neural networks that vary in their vulnerability to injury and capacity for repair and compensation. Early recovery may reflect resolution of transient physiological disturbances, whereas longer-term recovery depends more on structural repair, neuroplasticity, and functional network reorganization. Differences in white-matter damage, network redundancy, and compensatory recruitment may therefore produce function-specific recovery trajectories. In addition, recovery of higher-order functions may depend on restoration of more fundamental processes such as attention and processing speed. Another possible explanation is that TBI recovery is heavily influenced by initial injury severity, so it is possible that the stronger recovery of some measures we observed is due to our cohort having less severe TBI ^36–38^.

As discussed earlier, current literature suggests some cognitive impairment associated with TBI may be permanent ^31,32^. The tests for auditory encoding (HVLT Trials 1-3), auditory retrieval (HVLT Delayed Recall), and verbal processing speed (Stroop Word and Stroop Color) revealed significant differences between the TBI-18M and HC groups (Table 2), whereas there were no significant differences between TBI-18M and PTE. These findings imply that the impairment of these cognitive functions may be long-lasting or have a significantly slower recovery of function, which was not captured within the timeframe of this study. A slower recovery of functions is likely for auditory encoding and verbal processing speed, as the comparison between TBI-6M and TBI-18M showed a trend of recovery. This is supported by existing literature that verbal processing speed shows slower recovery post-TBI ^39^. In contrast, auditory retrieval appears to result in a more permanent loss of function, as evidenced by the similar performance levels observed in both TBI-6M vs. HC and TBI-18M vs. HC (Figure 1A). These four measures, with the exception of Stroop Color, showed a greater degree of significance between TBI-6M vs HC than between PTE vs HC, suggesting that either the initial TBI produces a stronger cognitive deficit in the associated functions than the second-hit from the transition to PTE ^9^, and/or the transition to PTE does not produce a significant cognitive deficit in these functions.

### Neurocognitive Profile of Post-Traumatic Epilepsy

When considering the findings from the PTE vs. HC and TBI-6M vs. HC comparisons, it becomes evident that while cognitive impairments resulting from a TBI exhibit deficits comparable in magnitude of functional impact to those seen in PTE (Table 2), the cognitive deficits associated with PTE are spread across a wider range of measures. Specifically, there are 11 out of 15 significant cognitive tests showing impairments in the PTE group compared to only 7 out of 15 in the TBI-6M group. To address the possibility of the PTE injury simply having a more pronounced impact on overall cognitive ability, rather than showing true isolated defects in cognition measures, we conducted a Spearman’s correlation to assess for redundancies among the cognitive measures. This revealed no significant correlations in performance between measures beyond those from the same original battery. Therefore, our increased number of significant measures in PTE are true findings and not multiple measures of a single construct. As there is an increase in the number of cognitive domains affected in PTE when compared to TBI alone, two possibilities are that the initial injury in PTE produced more widespread impairment and the impairment persisted, or that during recovery from the initial TBI, epileptogenesis lead to additional impairment.

Measures that assessed auditory attention span/capacity and working memory (i.e. Digits Forward and Digits Backward) showed no significant differences in performances between any cohorts or timepoints. Additionally, visuospatial encoding/retrieval (i.e. BVMT Trials 1-3, BVMT Delayed Recall) showed significance when comparing PTE vs HC, but did not show significance in any other cohort comparisons (Table 2). These results should be interpreted with caution, as the study may not have been sufficiently powered to show the significant difference in these cognitive domains.

### Potential Cognitive Markers of PTE

The significant difference observed between TBI-18M and PTE in written and oral processing speed (as measured by SDMT Written/Oral), as well as in visual working memory (evaluated with Dots), along with the lack of significant difference in the same tests between TBI-18M and HC, indicates that these tests reflect recovery in cognitive function following TBI (Table 2). Additionally, written/oral processing speed measures have been previously validated as highly sensitive measure for impaired processing speed ^40^. Since written and oral processing speed showed strong recovery trends after TBI and the absence of recovery in those who developed PTE, a slowed or lack of recovery in an individual suffering from TBI may be associated with the development of PTE.

Our theoretical domain analysis yielded results that mirrored the measure-level analysis, with Executive Function and Complex Processing Speed showing significant recovery of function in the TBI cohort (Figure 1B) and Auditory Learning and Memory showing chronic impairment vs prolonged recovery in TBI (Figure 1B). These domains can be utilized as simplified measures of cognition to monitor appropriate recovery following TBI. However, analyzing individual cognitive measures offers insights into specific impairments that may not be reflected in domain-level analyses. Our analysis indicates that while domain-based cognitive metrics can more reliably capture population-level cognitive outcomes, individual measure-based analysis has greater potential for identifying specific differences between cognitive outcomes related to TBI and PTE. This suggests that a detailed examination of individual measures might be more effective for uncovering potential biomarkers for PTE.

### Limitations and Future Directions

Potential limitations of this study, in order of importance, include brief follow-up period, selection bias, small sample size, lack of PTE development in the TBI cohort, and demographic and injury characteristic mismatch between cohorts. The follow-up period of 18 months may not be enough to fully characterize the recovery period post-TBI, such as we discussed with the results of the auditory encoding and retrieval. Selection bias likely affected our cohorts, as difficulty with activities of daily living (ADLs) may have prevented subjects with more severe interference related to their TBI from participating in the study and may have favored subjects who were naturally more predisposed towards recovery from TBI. Our cohort sizes were also limited, which may have masked significant differences in cognitive measures between cohorts, particularly when comparing TBI to PTE. Further subject-level analysis is needed to better understand which specific cognitive measure changes are more strongly implicated in the development of PTE, which also was complicated by the lack of longitudinal data within the PTE cohort. Furthermore, none of our current TBI cohort develop PTE during our study follow-up period. GCS scores were also unavailable for the PTE cohort. While injury symptomatic characteristics were used to assess for similar injury levels, GCS is still a strong predictor of post-traumatic seizures and worse cognitive/behavioral outcomes ^37,38^. Additionally, the control cohort tended to have a higher degree of education than the TBI or PTE cohorts, but this was adjusted for in the lme model.

## Conclusion

TBI subjects show an overall trend towards recovery in cognitive function which is evident from their trend in cognitive assessment measures tracked longitudinally post-TBI. Written/oral processing speed/working memory showed the strongest trend towards TBI recovery, while auditory encoding, auditory retrieval, and verbal processing speed showed chronic impairment. Stalled recovery, especially in oral/written processing speed and working memory may indicate a greater likelihood of developing PTE.

## Data Availability

All data produced in the present study are available upon reasonable request to the authors

---

**Supplemental Table 1.** Tbi Supplementary Data. Count data for causes of TBI and injury characteristics for the TBI and PTE cohorts. Dates that subjects sustained their initial TBI, as well as the time that has passed between initial TBI and testing sessions. Pearson correlation analysis was used to evaluate relationship between time between TBI and session 1 and cognitive measure performance, with no significance being found in any measures.

|  | PTE<br>(Y/N) | TBI<br>(Y/N) |
| --- | --- | --- |
| **Cause of TBI** |  |  |
| *Assault, attack, or other violence* | 3/17 | 3/29 |
| *Fall from dangerous height* | 2/18 | 8/24 |
| *Road, traffic, or other motor vehicle accident* | 5/25 | 13/19 |
| *Sports related injury* | 3/17 | 2/30 |
| *Other/not specified* | 7/13 | 6/26 |
| **Injury symptomatic characteristics** |  |  |
| *LOC <5 minutes* | 3/17 | 6/26 |
| *LOC >5 minutes* | 4/16 | 16/16 |
| *Amnesia < 30 minutes* | 1/19 | 3/29 |
| *Amnesia > 30 minutes* | 2/18 | 9/23 |
| *Retrograde amnesia* | 6/14 | 11/21 |
| *Anterograde amnesia* | 8/12 | 11/21 |
| **Time between TBI and testing session 1** |  |  |
| *Mean (days)* | 4621.5 ±<br>4509 | 105.1 ±<br>51.5 |
| *Maximum/Minimum (days)* | 14953 /<br>241 | 212 / 17 |
| **Time between TBI and testing session 2** |  |  |
| *Mean (days)* |  | 299.9 ±<br>68.3 |
| *Maximum/Minimum (days)* |  | 452 / 199 |
| **Time between TBI and testing session 3** |  |  |
| *Mean (days)* |  | 478.8 ±<br>70.4 |
| *Maximum/Minimum (days)* |  | 597 / 353 |
| **Time between testing session 1 and testing session 2** |  |  |
| *Mean (days)* |  | 189.6 ±<br>26.8 |
| *Maximum/Minimum (days)* |  | 240 / 126 |
| **Time between testing session 2 and testing session 3** |  |  |
| *Mean (days)* |  | 186.7 ±<br>38.6 |
| *Maximum/Minimum (days)* |  | 289 / 119 |

